# A Real-life Study of the Positive Response to DAA-based Therapies for Hepatitis C in Brazil

**DOI:** 10.1101/2020.10.06.20207274

**Authors:** Simone Monzani Vivaldini, Rachel Abraão Ribeiro, Gláucio Mosimann Júnior, Karen Cristine Tonini, Gerson Fernando Mendes Pereira, Wildo Navegantes de Araújo

**Affiliations:** Department of Chronic Conditions Diseases and other Sexually Transmitted Infections, Secretariat for Health Surveillance, Ministry of Health – Brasília/DF, Brazil; Center for Tropical Medicine, Faculty of Medicine, University of Brasília (UnB), Brasília/DF, Brazil; National Institute for Science and Technology for Health Technology Assessment, Porto Alegre/RS, Brazil; Faculty of Ceilândia, University of Brasília (UnB), Brasília/DF, Brazil

**Keywords:** hepatitis C, treatment, direct-acting antivirals, health policy, Brazil

## Abstract

Treating hepatitis C virus (HCV) infections reduces overall mortality and the risk of multiple extrahepatic complications. Direct-acting antivirals (DAAs) are molecules that target specific non-structural proteins of the virus, resulting in the disruption of viral replication and infection. This report describes the activities undertaken to assess the cure of HCV in patients of different age groups, supported by the Viral Hepatitis National Program of the Unified Health System in Brazil.

Around 11,000 patients wereevaluated in an electronic retrospective cohort, which used therapeutic schemes with Sofosbuvir (SOF), Daclatasvir (DCV), Simeprevir (SMV), and an association of Ombitasvir, Veruprevir/Ritonavir and Dasabuvir (3D) with or without Ribavirin (RBV), with sustained virologic response (SVR) or viral cure after a 12-week treatment. We conducted logistic regressions to identify factors independently associated with positive response to DAA-based therapies.

Among evaluated patients, 57.1% were male; 48.3% identified as white; 78.3% were over 50 years old; 44.1% were from the Southeast region; 47.7% had genotype 1b; and 84.5% were treated for 12 weeks. The SVR rates with DAAs ranged from 87% to 100%. Genotypes 1 and 4 had a better response (96.3 to 100% of SVR) to therapy with DAAs, and genotypes 2 and 3 had SVR of 90.6% to 92.2%. Different treatment periods showed SVR with an average of 95.0% and 95.9% for 12 and 24 weeks, respectively. In the odds ratio assessment, we verified that females were half as likely (OR 0.5; CI 95% 0.4-0.6) to have a negative response to therapy, when compared to males, and that genotypes 2 and 3 were around twice more likely (OR 1.5-2.2; CI 95% 0.7-2.9; 1.2-3.6 and OR 2.7-2.8; CI 95% 2.0-3.8, respectively) to not have SVR after therapy when compared to genotype 1. Patients aged between 50 and 69 years old were 1.2 times (OR 1,2; CI95% 0,7-1,9) more likely to not have SVR when treated with DAAs, when compared to other age groups.

This study is the first of this magnitude to be held in a Latin-American country with high SVR results, supported by a free-of-charge universal and public health system. We confirm the high performance of DAA-based therapies which support the Brazilian public health policy decision of adopting them as an important part of the country’s strategy to eliminate HCV by 2030.

## Introduction

Globally, there are an estimated 71 million people with chronic hepatitis C virus (HCV) infection and only a small minority were able to access curative treatment in 2016 ^[1, 2]^. Due to the substantial impact of hepatitis C virus (HCV) infection globally on patients, their families, and public health systems, the World Health Organization (WHO) has set HCV elimination goals that include the reduction of HCV incidence by 80% and HCV-related mortality by 65% before 2030 ^[1, 3]^. As part of the strategy to achieve these goals, WHO included all-oral, interferon-free, direct-acting antiviral (DAA) regimens in its 2017 edition ^[1, 3, 4]^.

About 632,000 people in Brazil are infected with HCV, with seroprevalence of 0.70 in the population aged between 15 and 69 years ^[5]^. Most HCV-infected people are unaware of their status until the virus has caused serious liver damage ^[3,6,7]^. The introduction of DAAs in 2015 has changed the landscape of treatment and its associated outcomes in the Brazilian Unified National Health System (SUS) ^[6]^. Despite the high potency of DAAs, their high cost initially represented a serious barrier, leading to the rationing of HCV treatment for patients with high risk of disease progression and the development of complications ^[7, 8, 9, 10, 11]^. However, today all Brazilian patients have access to these therapies.

Brazil’s National Report for HCV treatment was launched in 2015, elaborated by the Brazilian Ministry of Health (MoH-Brazil), and adopted by SUS ^[6]^. In 2018, the MoH-Brazil took a step forward to support the development and implementation of national multi-sector policies and strategies for hepatitis prevention and control in Brazil, for all HCV-infected patients, regardless of the degree of liver fibrosis ^[12]^. Since many countries are working on action plans to address HCV, different methodologies, processes, and ways to move forward have been identified. Therefore, the analysis and comparison of Brazil’s policies may offer an opportunity to accelerate the development and implementation of new national plans ^[13]^. SUS supported the annual cost per capita of US$ 6,152.85 on average for DAA-based therapies from 2015 to 2018 ^[14]^. The Ministry of Health has developed a mathematical model to better understand the national prevalence data and establish its own elimination goals, including an estimation of the number of people to be treated ^[5,15]^.

Since the isolation of the hepatitis C virus in 1989, several strategies to eliminate the infection have been studied. Interferon was the first drug used for treatment, having been used for many years. The addition of Ribavirin allowed the achievement of sustained virologic response (SVR) rates of around 55%. The most important change occurred with the introduction of DAAs: shorter treatments, oral use, and general SVR rates above 95% ^[15]^. The purpose of this study is to determine the impact of DAA regimens within all 27 Brazilian states and to identify the factors associated with SVR.

## Materials and Methods

A retrospective cohort study was conducted using different national databases of the Brazilian Public Health System, including the diagnostic information system (GAL), hospital information system (SIH), and treatment information system (HORUS). The population for this study included people with acute or chronic hepatitis C, most of them with important comorbidities. They were part of the first groups treated by the Brazilian National Health System (SUS) with therapeutic schemes which sometimes employed Ribavirin (RBV) and/or Peginteferon alfa-2a (PEG) but, in most cases, were composed of direct-acting antivirals: Sofosbuvir (SOF), Daclatasvir (DCV), Simeprevir (SMV) or Ombitasvir/Veruprevir/Ritonavir+Dasabuvir (3D). These drugs used different therapeutic combinations: SOF+DCV±RBV; SOF+SMV±RBV; SOF+RBV; 3D±RBV; SOF+PEG+RBV. Treatments conducted exclusively with Peginteferon alfa-2a and Ribavirin were also included with the objective of obtaining a comparative reference to DAA-based treatments.

To obtain the final database with the selected population, two steps were developed. The first step consisted of identifying a probabilistic relationship among the following databases: (i) Laboratory Environment Management System (GAL) – which contains data from the Central Laboratories of Public Health (LACEN); (ii) SUS Ambulatory Information System (SIA/SUS) – which includes records from the High Complexity Procedures Authorization (APAC) database; and (iii) Individual Outpatient Production Bulletins (BPAI) – which contain information recorded by states concerning requests and delivery of medication. From this relationship, a unique numeric code (ID) was attributed to each patient and this code was replicated in the different databases to allow the connection of the information taken from different sources. Later, patients’ personal data were suppressed to maintain anonymity. This step was developed by the SUS Department of Information Technology (DATASUS) through a project called “*VinculaSUS*”^[19]^, whose objective is to facilitate researchers’ analyses of policies developed by the Brazilian Ministry of Health while maintaining ethical commitments and the confidentiality of personal data.

The second step consisted of selecting patients with at least one viral load count by quantitative real-time qPCR-HCV test from the GAL database and some record of treatment for HCV in the SIA/SUS database, filtering patients who had detectable qPCR-HCV following 12 weeks of treatment and assessing how many of these patients presented SVR to HCV. Figure 1 details the procedures and quantifies the results of the first and second steps.

**Figure 1.**
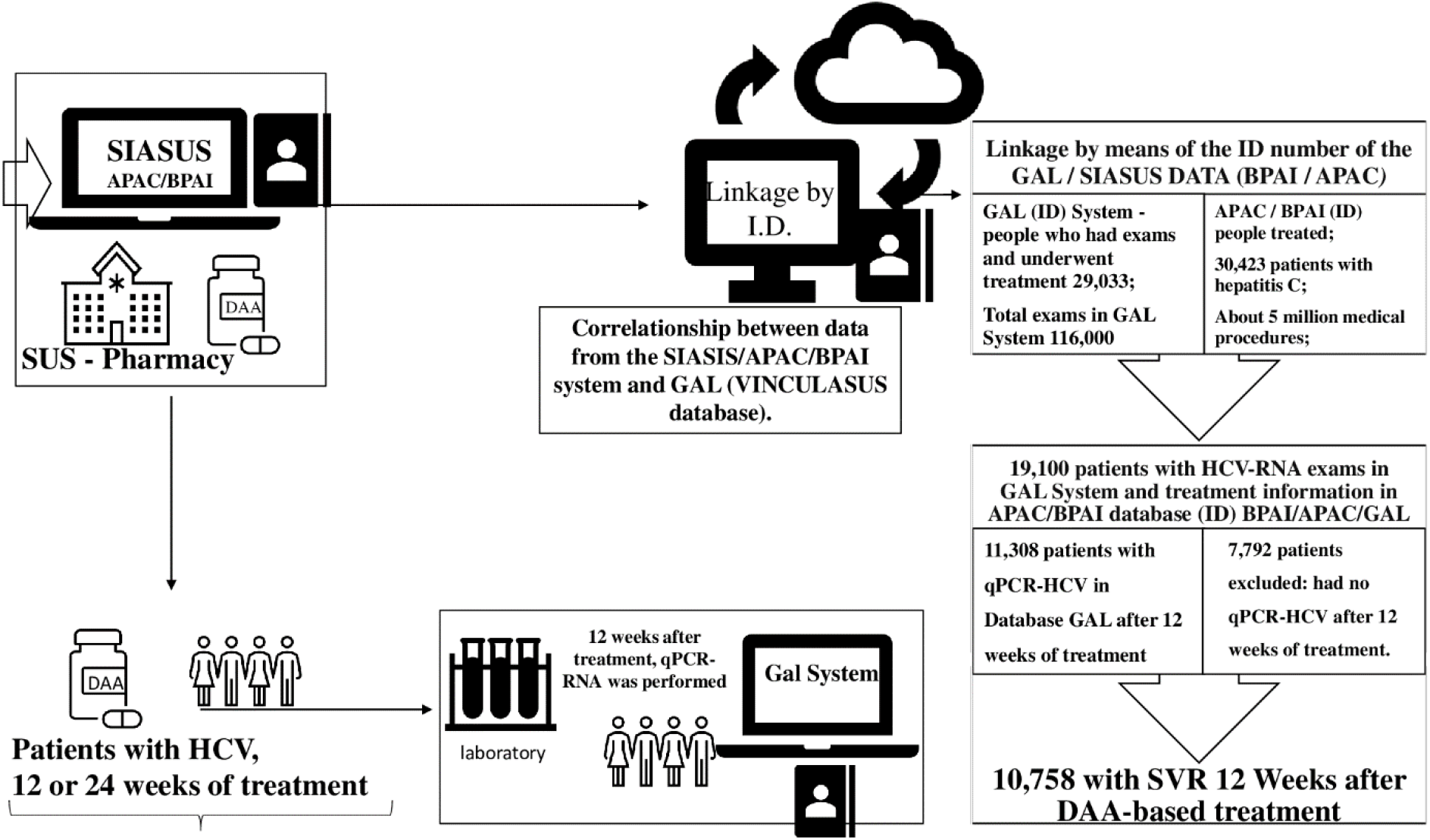
Relationship information from the databases used and final identification of 11,308 patients who completed the post-treatment investigation (quantitative qPCR-HCV test).

From the final database, descriptive analyses were performed with the objective of understanding the studied population and the procedures of interest that were adopted. All available sociodemographic, clinical, and laboratory variables were analyzed: sex, age, race/color, place of residence, genotype, time of treatment, therapeutic scheme, start and end date of treatment, date of laboratory tests, and respective results of qPCR-HCV tests. Trend and dispersion measures were used for numerical variables, and rates or proportions were utilized for categorical variables.

Patients were spatially distributed into three groups: 1) those registered in the APAC database who had at least one exam registered in the GAL database; 2) those registered in the APAC database who had at least one exam 12 weeks after the end of treatment registered in GAL; and 3) those registered in the APAC database who had at least one exam 12 weeks after the end of the treatment registered in GAL and who had reached SVR.

Some criteria were used to make the analyzed groups more homogeneous and to promote greater internal validity: analyses separated by genotypes; evaluation of therapeutic schemes; treatment time; and results of qPCR-HCV tests evaluated after 12 weeks of treatment completion. We compared the proportions to assess whether the results were statistically significant. Pearson’s chi-squared test was applied with a significance level of α = 5%. We used the odds ratio (OR) and confidence interval (CI) of 95% to identify the factors associated with SVR.

A bivariate analysis was conducted to assess the relationships between positive and negative responses to therapy and the available sociodemographic data, along with some of the comorbidities. For the multivariate analysis, we used the variables that presented a p-value < 0.2 in the bivariate analysis. We identified collinearity in two variables using a correlation matrix, and variables that presented R^2^ > 0.5 were selected. Three different models were evaluated through the Akaike Information Criterion (AIC), and a multivariate model of logistic regression was defined to determine the independent association between SVR responses to therapy (cure vs no cure). These responses were defined by the results of the qPCR-HCV test as undetectable (SVR) or detectable (not SVR).

All statistical analyses were conducted using SPSS 20.0 for Windows, with a significance level of 0.05 (Table 2a e 2b). Statistical analyses and graphical representations were performed using the IBM-PASW Statistics version 18 and Microsoft Excel® 2013 programs. Analysis and maps were developed utilizing RStudio, a free software program for an integrated development environment for R, version 1.2.5019-6 ^[20]^.

**Table 1.** Sociodemographic, clinical, and laboratory characteristics of patients who underwent hepatitis C treatment, from October 2015 to July 2018 in Brazil.

| Characteristics | Patients |  |
| --- | --- | --- |
|  | Total (N=11,308) |  |
|  | N | % |
| **Sex** |  |  |
| Male | 6459 | 57.1 |
| Female | 4847 | 42.9 |
| Ignored | 2 | 0.0 |
| **race / color** |  |  |
| White | 5464 | 48.3 |
| Black | 598 | 5.3 |
| Asian | 965 | 8.5 |
| Brown | 1217 | 10.8 |
| Indigenous | 5 | 0.0 |
| Ignored | 3059 | 27.1 |
| **age group** |  |  |
| 0 to 9 | 0 | 0.0 |
| 10 to 19 | 2 | 0.0 |
| 20 to 29 | 71 | 0.6 |
| 30 to 39 | 516 | 4.6 |
| 40 to 49 | 1884 | 16.7 |
| 50 to 59 | 3970 | 35.1 |
| 60 to 69 | 3522 | 31.1 |
| Above 70 | 1343 | 11.9 |
| **Region of Residence** |  |  |
| North | 617 | 5.5 |
| Northeast | 825 | 7.3 |
| Southeast | 4983 | 44.1 |
| South | 4393 | 38.8 |
| Center-West | 490 | 4.3 |
| **Genotypes** |  |  |
| Genotype 1 | 56 | 0.5 |
| Genotype 1a | 1674 | 14.8 |
| Genotype 1b | 5395 | 47.7 |
| Genotype 2 | 282 | 2.5 |
| Genotype 3 | 1934 | 17.1 |
| Genotype 4 | 42 | 0.4 |
| Mixed Genotypes | 12 | 0.1 |
| Unknown | 1913 | 16.9 |
| **Treatment Time** |  |  |
| 12 weeks | 9554 | 84.5 |
| 24 weeks | 1752 | 15.5 |
| Unknown | 2 | 0.0 |

**Table 2A.** Demographic, clinical, and laboratory characteristics of patients, and factors associated with SVR, bivariate and multivariate analyzes.

| SVR |  |  |  |  |  |  |  |  |  |  |
| --- | --- | --- | --- | --- | --- | --- | --- | --- | --- | --- |
| Characteristics | No |  | Yes |  | Bivariate analysis |  |  | Multivariate analysis |  |  |
|  | Total (N= 550) |  | Total (N=10.758) |  | Odds Ratio | Confidence Interval (CI) 95% | p-value | Odds Ratio | CI 95% | p-value |
|  | N | % | N | % |  |  |  |  |  |  |
| **Sex** |  |  |  |  |  |  |  |  |  |  |
| Male* | 399 | 6.2 | 6060 | 93.8 | - | - | - | - | - | - |
| Female | 151 | 3.1 | 4696 | 96.9 | 0.5 | (0.4 ; 0.6) | < 0.0001 | 0.5 | (0.4 ; 0.6) | < 0.0001 |
| Unknown | 0 | 0.0 | 2 | 100.0 | 0.0 | - | - | 0.0 | - | - |
| **Race / color** |  |  |  |  |  |  |  |  |  |  |
| White* | 285 | 5.2 | 5179 | 94.8 | - | - | - | - | - | - |
| Black | 23 | 3.8 | 575 | 96.2 | 0.7 | (0.5 ; 1.1) | 0.1492 | 0.8 | (0.5 ; 1.3) | 0.4599 |
| Asian | 50 | 5.2 | 915 | 94.8 | 1.0 | (0.7 ; 1.4) | 0.9644 | 1.0 | (0.7 ; 1.5) | 0.8774 |
| Brown | 60 | 4.9 | 1157 | 95.1 | 0.9 | (0.7 ; 1.3) | 0.6837 | 1.0 | (0.7 ; 1.4) | 0.8939 |
| Indigenous | 0 | 0.0 | 5 | 100.0 | 0.0 | - | 0.7350 | 0.0 | - | 0.9893 |
| Unknown | 132 | 4.3 | 2927 | 95.7 | 0.8 | (0.7 ; 1.0) | 0.0648 | 0.9 | (0.7 ; 1.1) | 0.2054 |
| **Age range** |  |  |  |  |  |  |  |  |  |  |
| 10 to 19 | 0 | 0.0 | 2 | 100.0 | 0.0 | - | 0.3587 | - | - | - |
| 20 to 29 | 1 | 1.4 | 70 | 98.6 | 0.3 | (0.0 ; 2.3) | 0.2503 | - | - | - |
| 30 to 39* | 23 | 4.5 | 493 | 95.5 | - | - | - | - | - | - |
| 40 to 49 | 83 | 4.4 | 1801 | 95.6 | 1.0 | (0.6 ; 1.6) | 0.9595 | - | - | - |
| 50 to 59 | 211 | 5.3 | 3759 | 94.7 | 1.2 | (0.8 ; 1.9) | 0.4105 | - | - | - |
| 60 to 69 | 182 | 5.2 | 3340 | 94.8 | 1.2 | (0.7 ; 1.8) | 0.4929 | - | - | - |
| Above 70 | 50 | 3.7 | 1293 | 96.3 | 0.8 | (0.5 ; 1.4) | 0.4660 | - | - | - |
| **Region of Residence** |  |  |  |  |  |  |  |  |  |  |
| **North\*** | 37 | 6.0 | 580 | 94.0 | - | - | - | - | - | - |
| **Northeast** | 48 | 5.8 | 777 | 94.2 | 1.0 | (0.6 ; 1.5) | 0.8867 | 0.9 | (0.5 ; 1.4) | 0.5730 |
| **Southeast** | 206 | 4.1 | 4777 | 95.9 | 0.7 | (0.5 ; 1.0) | 0.0332 | 0.8 | (0.6 ; 1.3) | 0.3997 |
| **South** | 243 | 5.5 | 4150 | 94.5 | 0.9 | (0.6 ; 1.3) | 0.6377 | 1.0 | (0.6 ; 1.4) | 0.8371 |
| **Center-West** | 16 | 3.3 | 474 | 96.7 | 0.5 | (0.3 ; 1.0) | 0.0372 | 0.7 | (0.4 ; 1.3) | 0.2121 |
| **Genotypes** |  |  |  |  |  |  |  |  |  |  |
| **1** | 0 | 0.0 | 56 | 100.0 | 0.0 | - | 0.3004 | 0.0 | - | 0.9666 |
| **1a\*** | 62 | 3.7 | 1612 | 96.3 | - | - | - | - | - | - |
| **1b** | 180 | 3.3 | 5215 | 96.7 | 0.9 | (0.7 ; 1.2) | 0.4705 | 0.9 | (0.6 ; 1.2) | 0.3538 |
| **2** | 22 | 7.8 | 260 | 92.2 | 2.2 | (1.2 ; 3.6) | 0.0022 | 1.5 | (0.7 ; 2.9) | 0.2831 |
| **3** | 181 | 9.4 | 1753 | 90.6 | 2.7 | (2.0 ; 3.6) | < 0.0001 | 2.8 | (2.0 ; 3.8) | < 0.0001 |
| **4** | 1 | 2.4 | 41 | 97.6 | 0.6 | (0.1 ; 4.7) | 0.6553 | 0.6 | (0.1 ; 4.5) | 0.6200 |
| **Mixed genotypes** | 0 | 0.0 | 12 | 100.0 | 0.0 | - | 0.9826 | 0.0 | - | 0.9844 |
| **Unknown** | 104 | 5.4 | 1809 | 94.6 | 1.5 | (1.1 ; 2.1) | 0.0143 | 1.2 | (0.9 ; 1.8) | 0.1968 |

**Table 2B.** Demographic, clinical, and laboratory characteristics of patients, and factors associated with SVR, bivariate and multivariate analyzes.

| SVR |  |  |  |  |  |  |  |  |  |  |
| --- | --- | --- | --- | --- | --- | --- | --- | --- | --- | --- |
| Characteristics | No |  | Yes |  | Bivariate analysis |  |  | Multivariate analysis |  |  |
|  | Total (N= 550) |  | Total (N=10.758) |  | Odds Ratio | Confidence Interval (CI) 95% | p-value | Odds Ratio | CI 95% | p-value |
|  | N | % | N | % |  |  |  |  |  |  |
| Therapeutic scheme |  |  |  |  |  |  |  |  |  |  |
| 3D + RBV | 0 | 0.0 | 30 | 100.0 | 0.0 | - | 0.4212 | 0.0 | - | 0.9757 |
| PEG + RBV | 43 | 27.6 | 113 | 72.4 | 7.4 | (5.1 ; 10.8) | < 0.0001 | 7.5 | (5.0 ; 11.2) | < 0.0001 |
| SOFO + DCV* | 199 | 4.9 | 3859 | 95.1 | - | - | - | - | - | - |
| SOF + DCV +RBV | 159 | 4.0 | 3835 | 96.0 | 0.8 | (0.6 ; 1.0) | 0.0449 | 0.8 | (0.6 ; 0.9) | 0.0094 |
| SOFO + PEG + RBV | 9 | 4.9 | 175 | 95.1 | 1.0 | (0.5 ; 2.0) | 0.9938 | 0.5 | (0.2 ; 1.0) | 0.0369 |
| SOF + RBV | 29 | 7.8 | 342 | 92.2 | 1.6 | (1.1 ; 2.5) | 0.0161 | 1.4 | (0.8 ; 2.5) | 0.2218 |
| SOF + SMV | 79 | 4.6 | 1649 | 95.4 | 0.9 | (0.7 ; 1.2) | 0.5888 | 1.3 | (1.0 ; 1.8) | 0.0484 |
| SOF + SMV + RBV | 13 | 3.3 | 377 | 96.7 | 0.7 | (0.4 ; 1.2) | 0.1671 | 1.0 | (0.5 ; 1.7) | 0.9092 |
| Unknown | 16 | 4.8 | 318 | 95.2 | 1.0 | (0.6 ; 1.6) | 0.9264 | 1.0 | (0.6 ; 1.8) | 0.8644 |
| Others | 3 | 4.8 | 60 | 95.2 | 1.0 | (0.3 ; 3.1) | 0.9587 | 1.2 | (0.4 ; 3.9) | 0.7585 |
| Time of treatment |  |  |  |  |  |  |  |  |  |  |
| 12 weeks* | 479 | 5.0 | 9075 | 95.0 | - | - | - | - | - | - |
| 24 weeks | 71 | 4.1 | 1681 | 95.9 | 0.8 | (0.6 ; 1.0) | 0.0862 | 0.8 | (0.6 ; 1.1) | 0.1973 |
| Unknown | 0 | 0.0 | 2 | 100.0 | 0.0 | - | 0.3904 | 0.0 | - | 0.9935 |
| Comorbidities |  |  |  |  |  |  |  |  |  |  |
| Neoplasm |  |  |  |  |  |  |  |  |  |  |
| Yes* | 48 | 8.0 | 551 | 92.0 | 1.8 | (1.3 ; 2.4) | 0.0003 | 1.7 | (1.2 ; 2.3) | 0.0015 |
| No | 502 | 4.7 | 10207 | 95.3 | 1.0 | (0.9 ; 1.1) | 1.0000 | 1.0 | (0.9 ; 1.1) | 1.0000 |
| HIV |  |  |  |  |  |  |  |  |  |  |
| Yes* | 70 | 4.6 | 1461 | 95.4 | - | - | - | - | - | - |
| No | 480 | 4.9 | 9299 | 95.1 | 1.1 | (0.8 ; 1.4) | 0.5695 | - | - | - |
| Fibrosis/Cirrhosis |  |  |  |  |  |  |  |  |  |  |
| Yes* | 171 | 4.6 | 3551 | 95.4 | - | - | - | - | - | - |
| No | 379 | 5.0 | 7209 | 95.0 | 1.1 | (0.9 ; 1.3) | 0.3524 | - | - | - |
| Child -Pugh A |  |  |  |  |  |  |  |  |  |  |
| Yes* | 1 | 16.7 | 5 | 83.3 | - | - | - | - | - | - |
| No | 549 | 4.9 | 10755 | 95.1 | 0.3 | (0.0 ; 2.2) | 0.2129 | - | - | - |
| Child-Pugh BC |  |  |  |  |  |  |  |  |  |  |
| Yes* | 1 | 3.3 | 29 | 96.7 | - | - | - | - | - | - |
| No | 549 | 4.9 | 10731 | 95.1 | 1.5 | (0.2 ; 10.9) | 0.6984 | - | - | - |
| Fibrosis |  |  |  |  |  |  |  |  |  |  |
| Yes* | 7 | 4.5 | 150 | 95.5 | - | - | - | - | - | - |
| No | 543 | 4.9 | 10610 | 95.1 | 1.1 | (0.5 ; 2.4) | 0.8126 | - | - | - |
| HBV |  |  |  |  |  |  |  |  |  |  |
| Yes* | 5 | 4.0 | 119 | 96.0 | - | - | - | - | - | - |
| No | 545 | 4.9 | 10641 | 95.1 | 1.2 | (0.5 ; 3.0) | 0.6659 | - | - | - |
| Diabetes |  |  |  |  |  |  |  |  |  |  |
| Yes* | 6 | 7.0 | 80 | 93.0 | - | - | - | - | - | - |
| No | 544 | 4.8 | 10680 | 95.2 | 0.7 | (0.3 ; 1.6) | 0.3633 | - | - | - |
| CKD |  |  |  |  |  |  |  |  |  |  |
| Yes* | 3 | 2.1 | 138 | 97.9 | - | - | - | - | - | - |
| No | 547 | 4.9 | 10622 | 95.1 | 2.4 | (0.8 ; 7.5) | 0.1406 | - | - | - |
| Portal hypertension |  |  |  |  |  |  |  |  |  |  |
| Yes* | 1 | 5.9 | 16 | 94.1 | - | - | - | - | - | - |
| No | 549 | 4.9 | 10744 | 95.1 | 0.8 | (0.1 ; 6.2) | 0.8452 | - | - | - |
| Transplanted |  |  |  |  |  |  |  |  |  |  |
| Yes* | 9 | 4.2 | 204 | 95.8 | - | - | - | - | - | - |
| No | 541 | 4.9 | 10556 | 95.1 | 1.2 | (0.6 ; 2.3) | 0.6626 | - | - | - |

As mentioned before, this study utilized a historical database, and participants’ names and personal information were kept confidential by substituting these data for an identification number (ID). As a result, the National Committee for Ethics in Research (CONEP) waived the requirement to obtain patients’ informed written consent and approved the research, which is registered in Brazil under number 2,872,543.

## Results

The SUS Department of Information Technology (DATASUS), through a project called VinculaSUS^[19]^, worked on establishing a relationship among the various Ministry of Health’s databases by providing a numeric identification key (ID) which enables locating patients in these various information systems and facilitates researchers’ analyses of policies adopted by the MoH-Brazil, while maintaining ethical commitments and the confidentiality of personal data. From this relationship, a unique numeric code (ID) was attributed to each patient and this code was replicated in the different databases to allow connecting the information taken from different sources.

During the analyzed period, 5 million ambulatory procedures were recorded in the SIA/SUS databases, including 30,423 patients treated for hepatitis C, of which 19,100 patients were registered with laboratory tests related to hepatitis C in the GAL database. Only 11,308 of these patients had a viral load test performed 12 weeks after the end of treatment, allowing the assessment of SVR and confirming the cure of hepatitis C. Of these 11,308 patients, 10,758 had a sustained virologic response and 550 individuals continued to present detectable qPCR-HCV following treatment (Figure 1).

When comparing proportions to assess whether the percentage differences per state among the three groups of patients were statistically significant, the states of the North, Northeast and Southeast regions presented the same proportion of cases (about 30%) in the three groups mentioned above (p-value >0.05). The other states showed significant percentage differences among the three groups (p-value < 0.05). This is due to the high concentration of patients in the first group from the Southeast region, primarily from the state of São Paulo, which represented 21.3% of the total number of patients. Although the other two groups represent the majority of cases in the Southeast region, the final analyzed group show a higher concentration of treatments for the states along the South-Southeast axis, translating into a more heterogeneous group.

Georeferencing of the treated and cured cases was carried out and can be seen in Figure 2, which shows the concentration of treatments in the South and Southeastern states.

**Figure 2.**
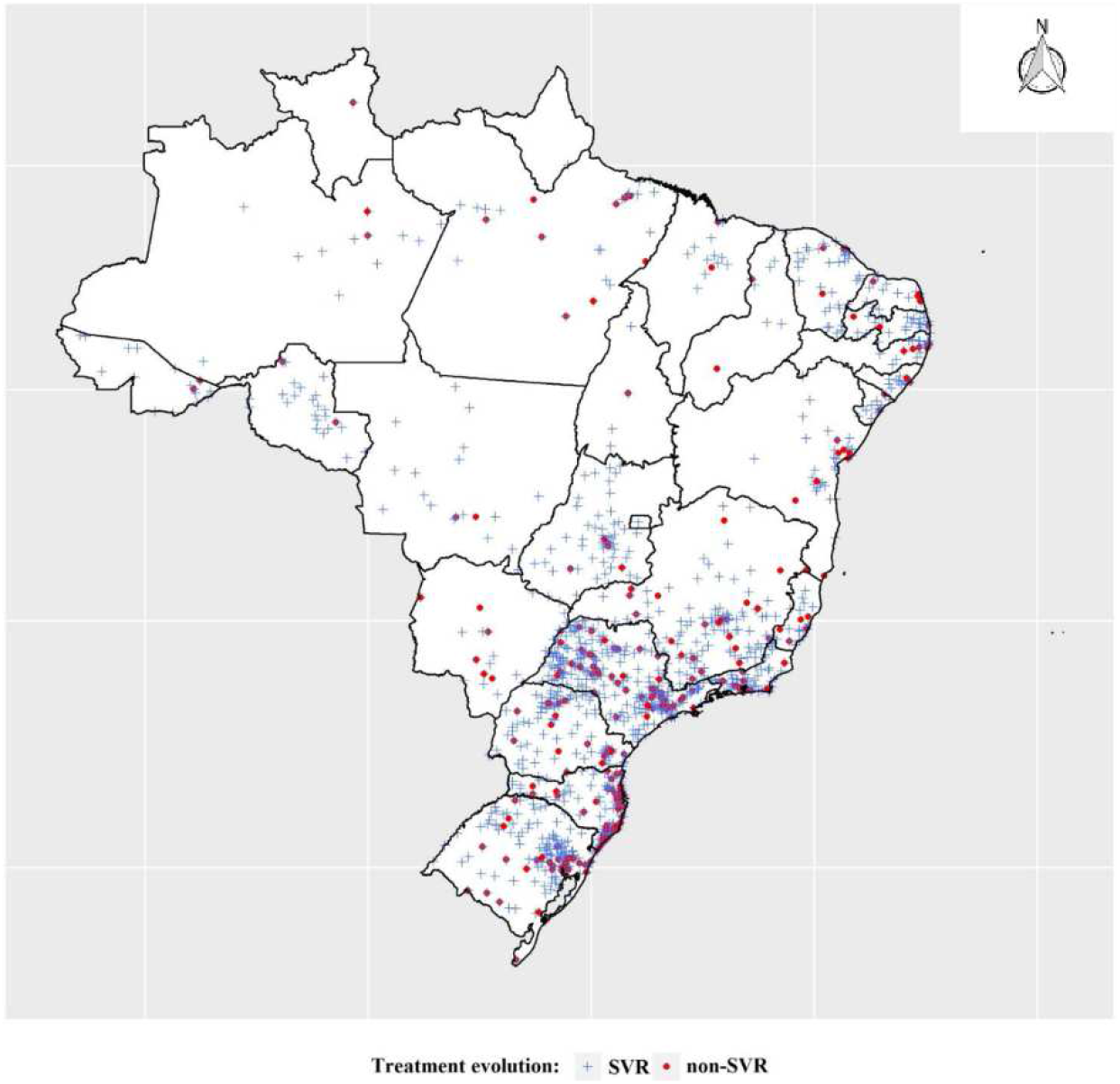
Georeferencing of cases according to patients treated for hepatitis C, and patients who achieved viral cure, Brazil, 2015 to 2018.

Among the evaluated patients (n = 11,308), 6,459 (57.1%) were male, 4,847 (42.9%) identified as white, 13,046 (65.8%) were between 50 and 69 years of age at the beginning of treatment, 8,686 (43.8%) lived in the Southeast region of the country, 4,983 (44.1%) had genotype 1b, 9,494 (84.0%), and 1,810 (16.0%) underwent treatment for 12 weeks and 24 weeks, respectively. More details are available in Table 1.

Among those with sustained virologic responses (SVR) or positive response to DAA with a undetectable qPCR-HCV at the end of treatment, 6,060 (56.3%) were male; 5,179 (48.1%) identified as white; 3,759 (34.9%) had initiated treatment at the age range of 50 to 59 years; 4,777 (44.4%) lived in the Southeast region; 5,215 (48.15%) carried genotype 1b; and 9,554 (85.5%) and 1,752 (15.5%) underwent treatment for 12 weeks and 24 weeks, respectively. Treatment regimens for each of the different genotypes identified in the study were evaluated to verify whether the different responses were statistically significant. Genotypes 1 (without subtype results), 1a, 1b, 2, 3, 4 and some mixed genotypes were present in the analyses. The average rate of SVR with the use of DAAs was 95.0% and 95.9% for 12 and 24 weeks after treatment, respectively (Table 2a).

In the odds ratio (OR) assessment, we verified that females are half as likely to have a negative response to therapy when compared to males (Table 2a).

In relation to the variable race/color, the rates did not present significantly different values for DAA-based therapies, with positive responses between 94.8 and 100%. The age group variable presented positive responses to treatment, varying from 94.7% to 100%; however, the p-value indicated no significant differences among the groups (Table 2a).

The positive response rate for the five Brazilian regions ranged from 94.0 to 96.7%, being significantly better in the Center-West and Southeast with an OR of 0.5 and 0.7, respectively (p< 0,05). SVR proportion ranged from 90.6% (genotype 3) to 100% (genotype 1). Some patients (12) were classified as infected by two different genotypes (mixed genotypes) and the SVR proportion was 100%. Patients with genotypes 2 and 3 obtained the lowest SVR proportion: 92.2% and 90.6%, respectively. When results were assessed according to genotypes, patients with genotypes 2 and 3 were 1.5 and 2.8 more likely to not have SVR, respectively, when compared to genotype 1a and 1b (Table 2b and Figure 3). These results were confirmed by the bivariate and multivariate analyses (Table 2a and 2b).

**Figure 3.**
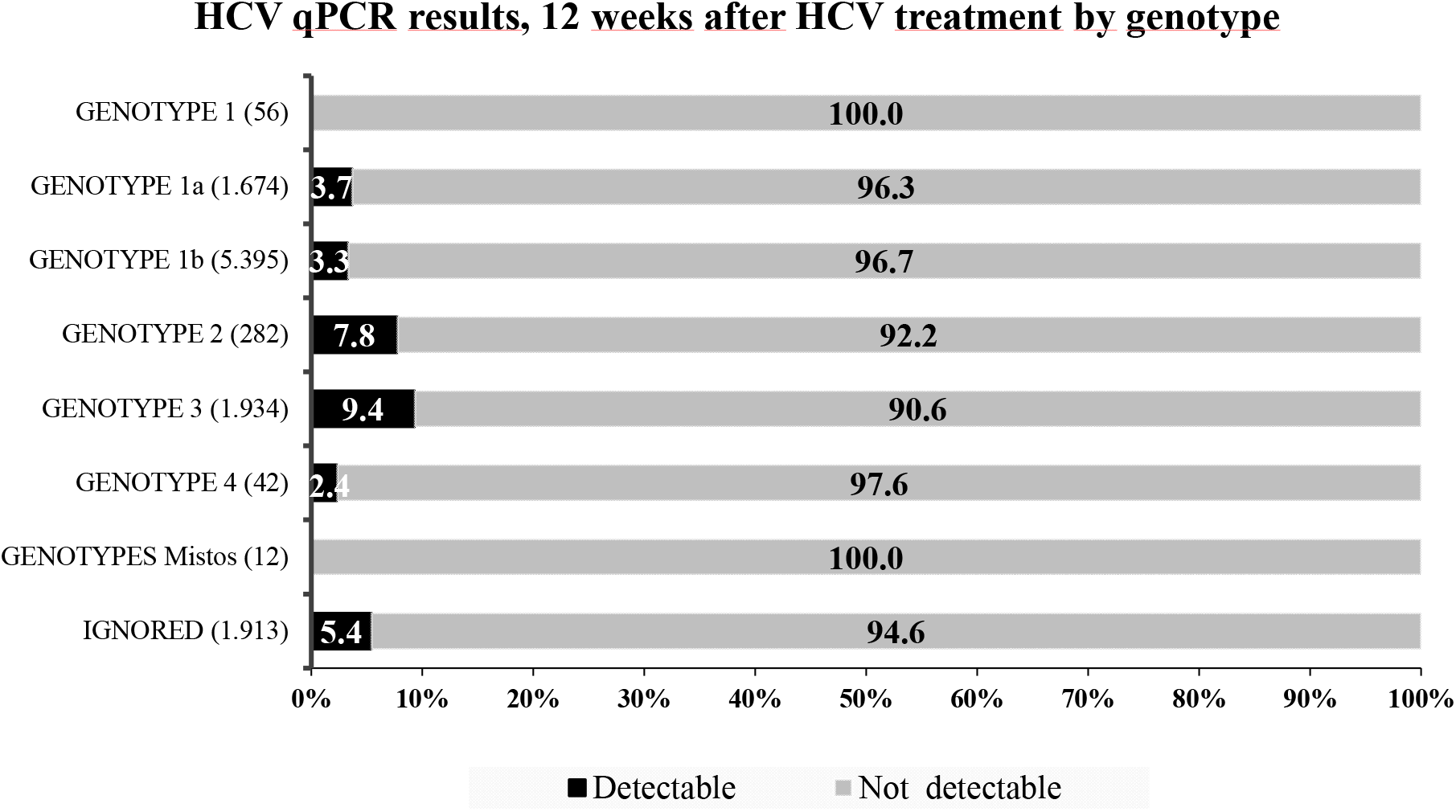
HCV qPCR results, 12 weeks after treatment for HCV, according to regimen and genotype, Brazil, 2015 to 2018.

The treatment of genotype 1a and 1b reached results of 96.3 and 96.7%, respectively. For 56 patients with genotype 1, there were no records of the subgenotype (1a and 1b). These patients had a 100% positive response rate to therapy (Table 2a and Figure 3).

In relation to the different therapeutic regimens assessed, the positive response rate or SVR ranged from 72.4 to 100%. The response rate for the PEG+RBV regimen was significantly negative when compared to the other schemes, with an OR of 7.5 (p<0.0001). The least efficient regimen was SOF+SMV, with an OR of 1.4. We found no differences between treatment time of 12 and 24 weeks (Table 2).

When observing the data by therapeutic scheme, the results of SVR after 12 weeks of DAA-based treatment ranged from 92.2% (Sofosbuvir + RBV) to 100% (3D + Ribavirin) (Table 2 and Figure 4)

**Figure 4.**
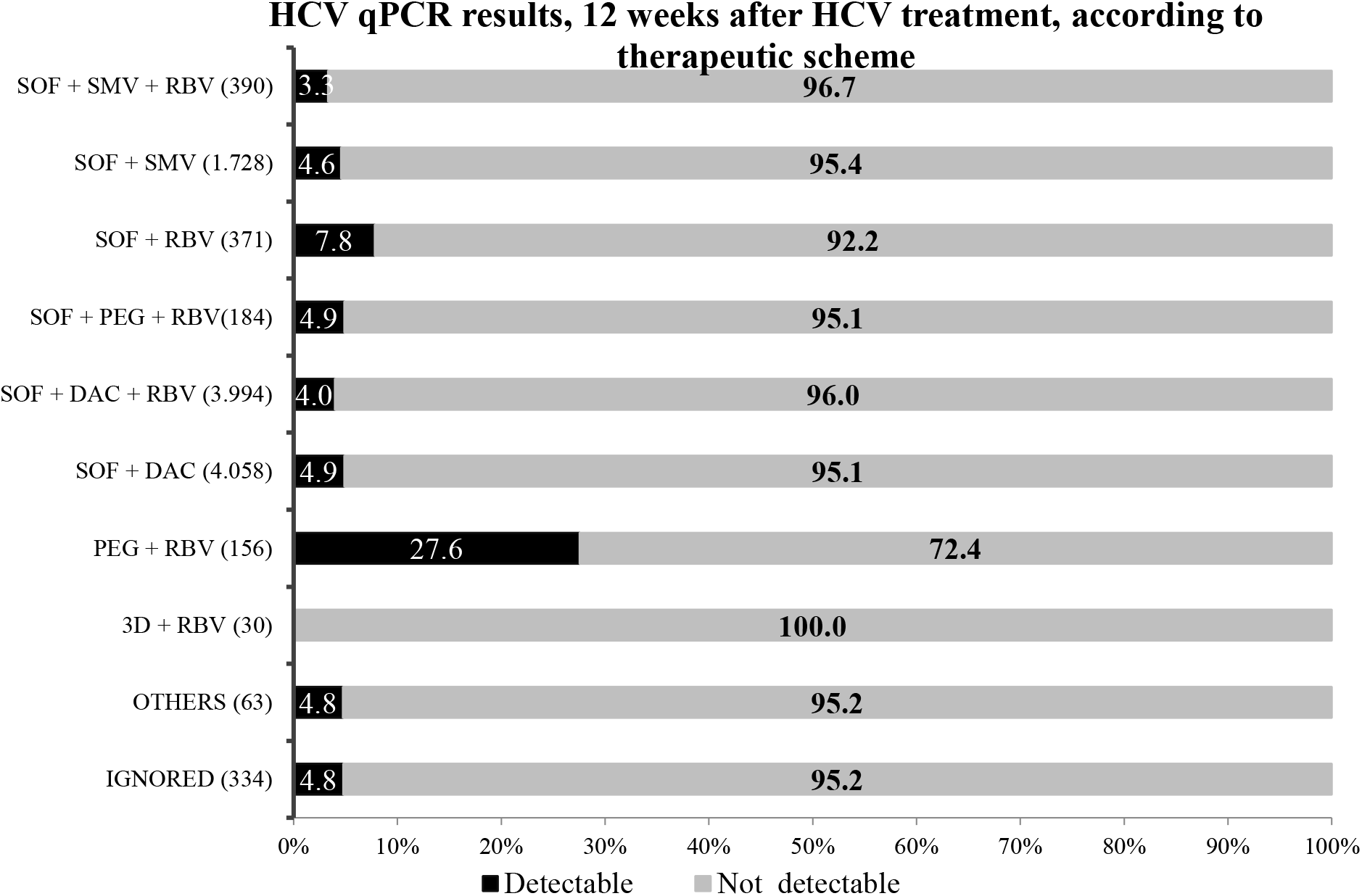
HCV qPCR results, 12 weeks after treatment for HCV, according to therapeutic scheme.

Patients with records of any of the comorbidities detailed in Table 2b had their information analyzed (7,588 registers). The response rate ranged from 83.3% (cirrhosis /child-Pugh A) to 97,9% (CKD-Chronic Kidney Disease); however, when the bivariate analysis was conducted, only the variable “presence of neoplasm” showed a significant result, with an OR of 1.8, that is, patients with neoplasms are nearly twice more likely to not reach a cure than patients without this comorbidity (p < 0,0003) (Table 2b).

Bivariate and multivariate analyses showed that genotypes 1a and 1b presented significantly different results when compared to others, responding better to therapeutic schemes. Genotypes 2 and 3, on the other hand, responded less to treatments when compared to genotype 1a and 1b. Genotype 4 had an SVR of 97.6%, but presented no significant difference due to the small number of cases. In the evaluation, it was observed that the SOF + RBV scheme had a significantly lower response to HCV (SVR 92.2%; OR 1.6; IC 95% 1,1-2.5) along with PEG + RBV (SVR 72.4%; OR 7.4; CI 5.1-10.8). We also verified that the SOF + DCV + RBV scheme yielded a higher response than the other antivirals (SVR 96%; OR 0.8; CI 0.6 -1.0). The SVR of 3D+RBV was 100% (Table 2b).

Table 3 shows the SVR rates according to the various therapeutic regimens and genotypes (1, 1a, 1b, 2, 3, and 4).

**Table 3.** HCV qPCR results following 12 weeks of treatment for hepatitis C for 11,308 patients in Brazil, according to the therapeutic scheme and patients’ genotype.

| Therapeutic scheme | Patients (N) | % | Genotypes | SVR |  |
| --- | --- | --- | --- | --- | --- |
|  |  |  |  | No (%) | Yes (%) |
| PEG+ RBV | 18 | 0.2% | genotype 1a | 38.9 | 61.1 |
|  | 40 | 0.4% | genotype 1b | 35 | 65 |
|  | 5 | 0.0% | genotype 2 | 40 | 60 |
|  | 25 | 0.2% | genotype 3 | 32 | 68 |
|  | 68 | 0.6% | ignored | 17.6 | 82.4 |
| 3D + RBV<br>with DAA | 4 | 0.0% | genotype 1a | - | 100 |
|  | 26 | 0.2% | genotype 1b | - | 100 |
| SOF+DAC | 18 | 0.2% | genotype 1 | - | 100 |
|  | 604 | 5.3% | genotype 1a | 3.5 | 96.5 |
|  | 1899 | 16.8% | genotype 1b | 2.5 | 97.5 |
|  | 20 | 0.2% | genotype 2 | 5 | 95 |
|  | 782 | 6.9% | genotype 3 | 11.9 | 88.1 |
|  | 31 | 0.3% | genotype 4 | 3.2 | 96.8 |
|  | 6 | 0.1% | Mixed genotypes | - | 100 |
|  | 698 | 6.2% | ignored | 5 | 95 |
| SOF+DAC+RBV | 18 | 0.2% | genotype 1 | - | 100 |
|  | 610 | 5.4% | genotype 1a | 2.3 | 97.7 |
|  | 1753 | 15.5% | genotype 1b | 2.7 | 97.3 |
|  | 9 | 0.1% | genotype 2 | - | 100 |
|  | 893 | 7.9% | genotype 3 | 7.3 | 92.7 |
|  | 8 | 0.1% | genotype 4 | - | 100 |
|  | 4 | 0.0% | Mixed genotypes | - | 100 |
|  | 699 | 6.2% | ignored | 4.6 | 95.4 |
| **SOF+PEG-RBV** | 8 | 0.1% | genotype 2 | 12.5 | 87.5 |
|  | 146 | 1.3% | genotype 3 | 4.1 | 95.9 |
|  | 30 | 0.3% | ignored | 6.7 | 93.3 |
| **SOF+RBV** | 14 | 0.1% | genotype 1a | 7.1 | 92.9 |
|  | 60 | 0.5% | genotype 1b | 3.3 | 96.7 |
|  | 225 | 2.0% | genotype 2 | 7.6 | 92.4 |
|  | 26 | 0.2% | genotype 3 | 11.5 | 88.5 |
|  | 46 | 0.4% | ignored | 13 | 87 |
| **SOF+SMV** | 10 | 0.1% | genotype 1 | - | 100 |
|  | 289 | 2.6% | genotype 1a | 5.5 | 94.5 |
|  | 1147 | 10.1% | genotype 1b | 4.4 | 95.6 |
|  | 3 | 0.0% | genotype 2 | - | 100 |
|  | 3 | 0.0% | genotype 3 | - | 100 |
|  | 2 | 0.0% | Mixed genotypes | - | 100 |
|  | 274 | 2.4% | ignored | 4.7 | 95.3 |
| **SOF+SMV+RBV** | 6 | 0.1% | genotype 1 | - | 100 |
|  | 84 | 0.7% | genotype 1a | 3.6 | 96.4 |
|  | 246 | 2.2% | genotype 1b | 4.1 | 95.9 |
|  | 54 | 0.5% | ignored | - | 100 |
| **unknown therapy** | 3 | 0.0% | genotype 1 | - | 100 |
|  | 36 | 0.3% | genotype 1a | - | 100 |
|  | 197 | 1.7% | genotype 1b | 4.1 | 95.9 |
|  | 11 | 0.1% | genotype 2 | - | 100 |
|  | 51 | 0.5% | genotype 3 | 11.8 | 88.2 |
|  | 1 | 0.0% | genotype 4 | - | 100 |
|  | 35 | 0.3% | ignored | 5.7 | 94.3 |
| **other therapies** | 1 | 0.0% | genotype 1 | - | 100 |
|  | 15 | 0.1% | genotype 1a | - | 100 |
|  | 27 | 0.2% | genotype 1b | - | 100 |
|  | 1 | 0.0% | genotype 2 | 100 |  |
|  | 8 | 0.1% | genotype 3 | - | 100 |
|  | 2 | 0.0% | genotype 4 | - | 100 |
|  | 9 | 0.1% | ignored | 22.2 | 77.8 |

When the results of treatments for each genotype were analyzed, we confirmed that the best SVR rates were reached by using SOF+DAC (+/-) RBV therapeutic regimens. The same was true for patients carrying genotypes 1a and 1b, whose SVR rates ranged from 96.5 to 97.7% (IC 95-98.2%).

## Discussion

About 11,308 patients had a viral load test performed 12 weeks after the end of treatment. The average rate of SVR with the use of DAAs was 95.0% and 95.9% for 12 and 24 weeks after treatment, respectively. It is important to highlight that the study cohort had a large number of patients with advanced fibrosis, cirrhosis, or comorbidities, which is consistent with the treatment protocol used from 2015 to 2018. The rates of SVR were very satisfactory, demonstrating that the treatment reached a high cure rate with an average of 95.5% with DAA incorporated into SUS (87% to 100%), being the largest study carried out to date in Latin America. Similar data were found in other studies ^[9,19,20]^.

Treatment schemes in each of the different genotypes identified in the study were evaluated to compare whether the different responses were statistically significant. Among those with sustained virologic responses (SVR), 56.3% were male; 48.1% identified as white; 34.9% had initiated treatment between 50 to 59 years of age; 44.4% lived in the Southeast region; 48.15% carried genotype 1b; and 85.5 % underwent treatment for 12 weeks. Genotypes 1 (without subtype results) 1a, 1b, 2, 3, 4 and some mixed genotypes were present in the analyses. Patients with genotypes 1a and 1b showed better cure rates than others (Table 2a). We also verified that treatments utilizing PEG + RBV resulted in an SVR of 72.4% and those that were DAA-based, such as 3D + RBV; SOFO + DCV +/-RBV; SOFO + PEG + RBV; SOF + RBV; SOF + SMV +/-RBV, obtained SVR rates ranging from 92.2 to 100%.

The analyses of the sociodemographic data showed that being a woman aged between 20 and 49 years and residing in the Southeast and Center-West regions were predictors of a better response to DAA-based therapies (Table 2a). The age range 50 to 69 years yielded a slightly lower response to DAAs when compared to other age groups (SVR of 94.8%), although this finding was not confirmed in the bivariate analysis, and the multivariate model could not be conducted (Table 2a). A large retrospective cohort study with about 17,500 patients with HCV, grouped into six age groups (<55 years of age, 55-59, 60-64, 65-69, 70-74, 75 years of age) and treated with DAA-based therapies in 2014/2015 found high SVR rates ranging from 90 to 94%, similar to those found in our study. It is worth highlighting that advanced age was not a predictor for a negative response, but rather the presence of cirrhosis. It is probable that in our cohort, people of more advanced age – being the largest group – had cirrhosis or neoplasms and that may have influenced the relatively lower SVR rates for this age group. However, when the factor age was analyzed independently, it was not a predictor of a lower SVR in relation to other age groups, according to other published studies ^[23–25]^.. The positive response rate in the five regions of the country ranged from 94.0 to 96.7%, being significantly better in the Center-West and Southeast, with an OR of 0.5 and 0.7, respectively. These finding are difficult to interpret, although quality of life and access to healthcare in these regions may contribute to a better regional response to DAAs. The state of São Paulo accounts for a large representation of patients in the Southeast region and this state is known as having a good public healthcare system. On the other hand, other conditions influencing the achievement or not of SVR are discussed in depth throughout this study.

The best therapeutic results for genotype 1a and 1b, utilizing these regimens, ranged from 96.3 to 96.7%, respectively. Patients with genotypes 2 and 3 obtained the lowest SVR rates (92.2% and 90.6%, respectively) and were 1.5 and 2.8 more likely to not have SVR, respectively, when compared to genotype 1a and 1b, a finding that has been confirmed in other studies ^[19, 20, 21, 22, 23, 24, 25]^.. Genotype 4 had an SVR of 97.6%, an excellent result, but the difference cannot be established as significant due to the small number of cases. Ferreira and collaborators evaluated 296 patients in the South region, obtaining SVR rates of 92% ^[16]^. Another study including 1,002 patients treated for HCV, with genotypes 1, 2 and 3, also carried out in the South region by Holzmann et al, found that even patients with liver cirrhosis achieved SVR rates ranging from 88.5% to 100% ^[27]^.In another study, in which 219 patients underwent therapy with DAAs, a high rate of SVR and excellent drug tolerability were found. Failure to achieve SVR was observed mainly among patients with at least one negative response predictor: cirrhosis and/or genotypes 2 or 3 ^[16]^. Other studies carried out in the Southeast region yielded similar data ^[18,19,20, 21,29,30]^.

This study showed that the SOF + RBV scheme had a significantly lower response to HCV (SVR 92.2%; OR 1.6; CI 95% 1.1-2.5) along with PEG + RBV (SVR 72.4%; OR 7.4; CI 5.1-10.8). We also verified a higher response for the SOF + DCV + RBV scheme than for other antivirals (SVR 96%; OR 0.8; CI 0.6 -1.0). The SOF+SMV regimen, with an SVR of 95.6%, yielded values that were very similar to SOF+DCV (95.1%); The addition of RBV to both schemes increased the rates to 96.7% and 96%, respectively (Table 2b). However, in the multivariate analyses, the SOF+SMV scheme proved to be less effective than SOF+DCV (OR 1.3), results that have also been corroborated in other studies ^[31,32]^.

Although the SVR of 3D + RBV was 100%, we were not able to verify whether this result was significant due to the small number of treated patients (Table 2b).

The high cure rate after using the therapeutic schemes considered in this study, with data from all Brazilian states, has great relevance to demonstrate that public policies developed by the Brazilian Ministry of Health, including the incorporation of therapies requiring large investments, providing cure for thousands of hepatitis C patients, has brought Brazil closer to the goal of eliminating the disease as a public health hazard, and over 140,000 people have been treated to date. This study can help to understand the dynamics of response to therapy in the Brazilian population. The average positive response with the use of DAAs ranged from 95.0% to 95.9%, demonstrating that it is possible to obtain good results with therapies for hepatitis C in the public health system.

### Limitations

This study was conducted using, initially, secondary data collected from the Ministry of Health’s information systems (DATASUS). However, in order to collect data, it was necessary to work with several independent areas of that institution, as there is no interface between the different systems. Therefore, investments to increase the performance of the Ministry’s systems – in charge of so many diseases – would facilitate research and permit more efficient analyses of data by managers at all levels (municipalities, states, and the federal government).

Although around 80,000 treatments were distributed throughout the period of this study, when the databases were merged, only 29,033 patients were found in the GAL database for undergoing some hepatitis C exam while also being registered in other databases related to the treatment of Hepatitis C (SIASUS - BPAI and APAC). Of these, 19,100 (66%) had qPCR-HCV in the GAL, but only 11,308 (∼60%) had qPCR-HCV after 12 weeks of treatment for hepatitis C (Figure 2). Therefore, it can be inferred that most patients may have undergone testing through Supplementary Health Care (Health Insurance Plans) or bore the costs of laboratory tests themselves, and therefore, were not included in the GAL system database.

Among the main limitations to this study, the incompleteness of data in all systems stands out. The main data refer toresults of viral genotype; race/color; and comorbidities (Tables 1 and 2). The study was conducted with secondary data that did not always adequately control confounding factors. The absence of sociodemographic data (resulting from poorly filled out forms), such as information about race/color (27%) is common in the various SUS databases (e,g.: GAL and Sinan-Notifiable Diseases Information System).

In the evaluated age groups, we found some differences in the SVR rate, but this was not confirmed by the bivariate analyses. SVR rates varied from 98.6 to 100% in the age group between 10 and 29 years. This indicates that there is a better response to therapy among younger populations; however, the number of patients assessed was small (73) and this positive difference in SVR could not be confirmed in the statistical models.

In relation to comorbidities, there were 7,588 registers; for the remaining patients (3,720), this variable had been left blank (absent information), a fact that may cause a bias of information as this information was absent for about 30% of the patients in this cohort. It is true that the guidelines in that period restricted SUS treatment for patients with hepatitis C who had advanced fibrosis (stages 3, 4) or those who presented some type of comorbidity. Although some comorbidities are known for their negative response to therapies, such as the presence of decompensated cirrhosis, the assessed data did not show significant differences in the SVR. This could be intrinsically related to incorrect or absent data, which may interfere in the analyses and increase bias, as mentioned above. Data related to comorbidities are exclusively filled out in spreadsheets and sent to the Ministry of Health; they are not included in the SIASUS system as are the others.

The only comorbidity that yielded a significantly negative difference when compared to the others (Table 2), was the presence of neoplasms (SVR of 92%; OR-1,8; p-value <0,0003). This result for people with some types of neoplasm was similar to the findings in another study ^[33]^. It is important to note that, even with a poorer result when compared to others, the SVR rates are still excellent and the benefit of treating people with neoplasms is positively reported in literature, with a rate of 92% viewed as very satisfactory. ^[24,25,34]^.

## Conclusion

Even without obtaining data for all patients initially located in the SIA/SUS, it is possible to conclude that the data in this study obtained excellent results, consistent with the study performed. Brazil is also a signatory to the World Health Organization resolution and has a National Plan for the Elimination of Hepatitis C as a public health hazard by 2030. One of the main goals of this plan is to offer more diagnosis and treatment for 50,000 people annually, by 2025 ^[13, 36]^. Our work shows that the incorporation of DAAs was of prime importance to achieve high cure rates in the studied population and highlights the importance of monitoring treatments and achieving virologic cure. The Brazilian Public Health System must be included among the public institutions that offer treatments to its population in a universal and egalitarian way.

Currently, new treatments are being incorporated into SUS, with other drugs that are as good or better than the ones we studied. Our study demonstrates that the excellent results obtained in curing patients with hepatitis C may have even better results in the future. This is significantly important, confirming the excellent results of the treatment of hepatitis C in the national territory and the need to maintain treatment for the elimination of hepatitis C as a public health problem by 2030 ^[5,14,36]^.

## Data Availability

As mentioned before, this study utilized a historical database, and participants names and personal information were kept confidential by substituting these data for an identification number (ID). As a result, the National Committee for Ethics in Research (CONEP) waived the requirement to obtain patients' informed written consent and approved the research, which is registered in Brazil under number 2,872,543.

## Acknowledgements

We would like to acknowledge all of the support provided in the preparation and publication of this study, Brazil’s Ministry of Health, DATASUS and the Department of Chronic Conditions Diseases, and other Sexually Transmitted Infections/the Ministry of Health, teams who made this study possible.

## Authors’ contributions

**Conceptualization**: SMV, WNA; **Data search**: SMV, RAR, WNA; **Investigation**: SMV, RAR, WNA; **Project administration**: SMV, WNA; **Methodology:** SMV, WNA, RAR, **Supervision**: SMV, WNA; **Validation**: SMV, RAR, GMJr, KCT, GFMP**; Writing:** SMV, RAR, GMJr, GFMP, KCT; WNA; **Writing-review & editing:** SMV, RAR, GFMP, GMJr, KCT, WNA

